# Features Influencing Diagnostic Yield of Exome Sequencing in the DECIPHERD Study in Chile

**DOI:** 10.64898/2026.02.12.26345769

**Authors:** Gabriela Moreno, Boris Rebolledo-Jaramillo, Daniela Böhme, Gonzalo Encina, Luz María Martin, M. Jesús Zavala, Fernanda Espinosa, M. Trinidad Hasbún, M. Cecilia Poli, Víctor Faundes, Gabriela M. Repetto

## Abstract

**Background:** Exome sequencing (ES) has become a key diagnostic tool for rare diseases (RDs). However, most evidence on ES performance comes from high-income countries and patients from European ancestry. In countries such as Chile, limited access to next generation sequencing amplifies health disparities and highlights the need to identify which patients are most likely to benefit from ES.

**Methods:** This study presents the second phase of the Chilean DECIPHERD project, in which we performed ES in a new group of patients with RDs presenting with multiple congenital anomalies (MCA), neurodevelopmental disorders (NDD), and/or suspected inborn errors of immunity. To identify clinical and demographic factors associated with an increased probability of obtaining an informative ES result, we conducted a logistic regression analysis, combining the results of the first and second phases of the project. We also objectively evaluated global ancestry measured using ADMIXTURE, as a potential factor.

**Results:** Sixty-seven patients participated in this second phase of DECIPHERD with a median age of 6 years (range: 0–27); 55.2% were female, with an average (± s.d.) proportion of Native American ancestry of 0.615 ± 0.18. Clinically, 52.2% presented with both MCA and NDD, and the rest had other phenotype combinations. An informative result, including pathogenic or likely pathogenic variants in genes consistent with the patient’s phenotype, was identified in 34.3% of the cohort; 61% of these variants had not been previously reported in databases such as ClinVar. By combining the two phases of the study, we reached a total of 167 patients, in whom the presence of NDD and/or MCA significantly increased the probability of achieving an informative ES outcome. In contrast, previous use of gene panel testing was associated with a decreased likelihood of receiving an informative result. Ancestry was not associated with diagnostic yield.

**Conclusions:** This study demonstrates the utility of ES in achieving a diagnosis in a clinically diverse cohort of Chilean patients with RDs, and characterized features associated with a higher diagnostic yield. These findings may contribute to evidence-based patient prioritization strategies in settings with limited access to NGS resources.

## Introduction

Rare Diseases (RDs) are a global health concern, affecting millions of patients and marked by numerous challenges, including prolonged diagnostic delays. Given the genetic basis underlying many RDs, next generation sequencing (NGS) is utilized in both clinical and research settings, allowing thousands of patients from diverse cohorts to finally achieve a diagnosis (1–4) This improves access to information and clinical management of patients, caregivers and clinicians, in addition to contributing to new disease insights (5–8).

Prior research has demonstrated the utility of early use of exome sequencing (ES) and genome sequencing (GS) by NGS in the search for answers in patients with RDs. A pooled meta-analysis reported an overall diagnostic yield of 38% for ES (1); however, there is high variability in diagnostic yields of ES and GS across different clinical cohorts, ranging from 4-83% (1,2).

In line with this variability, several studies report that certain clinical features are associated with differences in diagnostic yield. For instance, cases with syndromic involvement and suspected metabolic disorders showed higher diagnostic yields reaching 43.8% and 64.3% respectively, than the overall study rate of 28.3% (9). Similarly, in patients presenting with neurodevelopmental disorders (NDD), the diagnostic yield was higher (43.0%) than the overall cohort that included other phenotypes (29.3%) (4). Wright et al (3) reported an increased chance of receiving a diagnosis for patients presenting severe intellectual disability or developmental delay (OR = 2.41). Conversely, lower diagnostic yield has been associated with extreme prematurity (OR = 0.39) and African ancestry (OR= 0.51) (3). Similar findings have been reported in a GS study of 822 families with diverse phenotypes, in which diagnostic yields were lower among non-European ancestries, including families of Admixed American ancestry, who achieved a diagnostic yield of 15% compared with the overall diagnostic yield of 29.3% (4). This suggests that guidelines for selection of patients with higher likelihood of diagnosis may be helpful to steer clinical decision-making, especially in settings with limited resources.

In Latin America in general, and in Chile in particular, access to NGS in clinical settings is limited, given the lack of local capacities and financial coverage by health systems (10–12). These limitations leave many patients undiagnosed. In high-income countries (HICs) that have incorporated ES and GS in their health systems, an undiagnosed rare disease (URD) usually reflects the current technical limits of genomic medicine and knowledge of disease mechanisms, such as gaps in short-read sequencing or incomplete gene–phenotype understanding, because patients typically have timely access to exome/genome sequencing (13–15). In contrast, in low- and middle-income countries (LMICs) or even in HIC that do not have ES or GS incorporated as a health service, such as the current situation in Chile, the status of URDs more often stems from structural barriers. These includes limited availability of NGS, lack of local sequencing capacity, inadequate reimbursement, and reliance on lower-yield tests such as karyotype, Multiplex Ligation-dependent Probe Amplification**®** (MLPA), and NGS-based panels (Encina et al., 2019; Wainstock & Katz, 2023). Thus, in resource limited genomic settings, it is key for policy makers and clinicians to build local evidence to assist in identifying patients with higher likelihood of benefiting from ES testing, from a diagnostic yield, clinical utility and cost-effectiveness perspectives.

In Chile, ES is not routinely included in clinical assessments, and its performance, as well as the factors influencing the likelihood of achieving a diagnosis through ES in our population, remain unknown. Thus, this study has two main aims. First, we expand on the results of the Chilean <u>DE</u>coding <u>C</u>omplex Inherited <u>Phe</u>notypes in <u>R</u>are <u>D</u>isorders (DECIPHERD) project (16) presenting results for additional patients. Second, we assess the performance of ES for RDs in the Chilean population and identify factors that influence the likelihood of reaching a diagnosis, performing a joint statistical analysis of the results of all cases.

## Methods

### Participants

Individuals of any age with RDs of probable genetic etiology and their parents were recruited from tertiary healthcare facilities in Chile between January 2023 and December 2024 (second phase of the DECIPHERD project). They were eligible for the study if they had at least one of the three following clinical criteria: (1) multiple congenital anomalies (MCA), (2) neurodevelopmental disorders (NDD) and/or (3) suspected inborn errors of immunity (IEI) and had already undergone clinically available diagnostic evaluations without reaching a definitive causal or etiologic diagnosis. Inclusion criteria were the same as for the first phase of the DECIPHERD project (16).

### Procedures

Managing physicians including neurologists, geneticists, immunologists, and others, presented patient cases during weekly meetings to the DECIPHERD team; presentations followed a semi-structured methodology covering clinical manifestations, family health history, and prior test results. This information was described using Human Phenotype Ontology (HPO) terms, and patients’ phenotypes were grouped in systems, following HPO’s hierarchy (17). Clinical, phenotypic, and relevant variant information were stored in REDCap**®** (18).

If no evident diagnosis was made or suggested after these presentations, the clinical information was considered complete, and no further specific clinical testing was recommended, families were invited to participate. When accepted, blood samples from the proband and both parents (if available) were collected from a peripheral vein in EDTA tubes. When mosaic conditions were suspected, additional buccal swabs were obtained from the probands.

### Bioethics

This project was reviewed by the Institutional Ethics Review Board at Facultad de Medicina Clínica Alemana Universidad del Desarrollo (Approval ID 2018-045). All families provided written informed consent for participation and publication.

### Exome Sequencing

DNA samples were sent to Sistemas Genómicos© (Valencia, Spain) for exome sequencing. If a mitochondrial condition was suspected, sequencing of the mitochondrial DNA (mtDNA) was included. The number of interrogated genes was 19,123 or 19,160 in the latter case.

### Variant prioritization

Data from the second phase were analyzed on Sistemas Genómicos’ GeneSystems platform (GeneBytes© 6/19/2023 – 3/17/2024 or Genebytes X © 6/29/2024 – 3/18/2025). For each family, a list of HPO terms describing the proband’s phenotype was added to the analysis pipeline. Matching HPO terms were used as a prioritizing feature, not as a filter. Filters for the evaluation of SNVs were depth ≥ 10x, allelic fraction ≥ 0.2, population frequency (gnomAD and 1000 Genomes Project) ≤ 0.005, and consequence type: high (the variant has a major functional consequence, e.g. stop gain, frameshift, splice variant) or moderate (the variant likely changes the protein sequence, but may not completely abolish its function, e.g. missense variant, inframe variation). In addition, we analyzed CNVs considering the following filters available on the Sistemas Genómicos’© platform: bins ≥ 6 and Cnid Score ≥ 5. Duplications and deletions were searched against the ClinGen’s dosage sensitivity curations (19) and the DECIPHER database (20) to assess pathogenicity. Additionally, a manual literature search was conducted for the prioritized variants.

### Clinical variant interpretation

We defined two types of results: informative and non-informative. For the informative class we defined two subcategories: (*1*) *categorical*, if a variant was interpreted as pathogenic (P) or likely pathogenic (LP), in a gene associated with a concordant phenotype and mode of inheritance for the patient; and *(2) candidate*, if variants of uncertain significance (VUS) were identified in a gene associated with a concordant phenotype and mode of inheritance. “Non-informative” was considered in cases in which no variants were identified that could explain the patient’s phenotype. Patients with a single P/LP variant in a gene causing a recessive phenotype were considered as “special considerations” in the non-informative category.

Results were initially categorized during the analysis period of June 2023 to April 2025, and variant classification was subsequently updated in October 2025 in search for potential reclassifications using Franklin (21) and VarSome (22) platforms. When appropriate, additional ACMG criteria were applied manually to the automated classification provided by these platforms, including PP3, for computational evidence supporting a deleterious effect on the gene or gene product; PP4, for highly specific patient or family phenotypes; PM3 for variants associated with recessive disorders detected in trans with a pathogenic variant; and PS2, for *de novo* variants (with both maternity and paternity confirmed). When interpretations were discrepant, we analyzed the reasons for these discrepancies considering the ACMG/AMP points-based classification system (23), and kept the variant classification with more evidence.

### Ancestry analysis

For this purpose, patient VCF files were consolidated and merged with reference panels from the 1000 Genomes Project Phase 3, including European, African, and Admixed American samples (<90% admixture, considered as Native American). We applied ADMIXTURE (24) with k = 3 to calculate global ancestry components and subsequently compared Native American proportions between informative and not-informative patients (see below).

### Statistics

Descriptive analyses comprised the calculation of frequencies, proportions, percentages, means, medians, and ranges. The chi-square test was used to compare diagnostic rates across variables. t-test was used to compare the means of ancestry estimates. Logistic regression (bivariate and multivariate) was used to assess factors influencing the diagnostic yield. We considered a p-value < 0.05 as statistically significant. Statistical analyses and plots were generated using R (25).

To explore other factors that contributed to the likelihood of obtaining an informative result, we added the 103 cases from our previously published DECIPHERD first phase that had the same inclusion criteria and diagnostic categories (16). This resulted in a combined dataset of 167 cases. Three cases from the first phase were also included in the second phase because they were re-analyzed using a different sequencing strategy; for the regression model analyses, we retained the most recent results.

Using the combined dataset, we first constructed separate bivariate logistic regression models for each of the following variables: sex, age, the presence or absence of MCA, NDD and IEI, coexistence of different inclusion criteria (e.g., NDD and MCA) and number of affected systems. We also included variables related to prior testing and analytical approach, specifically whether a previous clinical gene panel has been performed and the family structure available for analysis (solo, duo or trio). Then, we developed a multivariate logistic regression model incorporating all variables that were statistically significant in the bivariate analysis adjusted by age and sex. Diagnostic rate was calculated as number and percentage of informative (categorical and candidate) among all reviewed cases.

## Results

### Participants: demographics and clinical features of the second phase

Seventy-eight cases were screened during the study period, 73 (93.6%) were invited to participate, but six were not enrolled: two declined to participate, three did not complete the enrollment process (informed consent signature or sample collection), and one was referred to another study. Sixty-seven patients (55.2% female) and their parents completed participation in the study period. Fifty-eight cases (86.6%) were analyzed as trios: 57 included the patient and both parents, and one included the patient, the mother, and an affected sister. The remaining nine cases (13.4%) were analyzed as duos: eight maternal and one paternal. In total, 192 individuals underwent ES.

The mean age of the mothers at the time of delivery was 28.3 ± 6.9 years (mean ± s.d.), while the mean age of the fathers was 33.0 ± 9.3 years. Demographic information about the patients is presented in Table 1.

**Table 1.** Demographic characteristics of the probands of the second phase of DECIPHERD.

|  |  | Age (yrs) at recruitment | Health care provider located in Santiago | Public health insurance |
| --- | --- | --- | --- | --- |
|  | N (%) | median [range] | N (%) | N (%) |
| **Female** | 37 (55.2) | 8 [0-27] | 31 (83.8) | 31 (83.8) |
| **Male** | 30 (44.8) | 5.5 [0-22] | 22 (73.3) | 27 (90.0) |
| **Total** | 67 (100) | 6 [0-27] | 53 (79.1) | 58 (86.6) |

In total, 64 cases (95.5%) in the second phase had undergone at least one previous non-informative genetic test (median: 2; range: 1-4). Specifically, 43 had a G-banded karyotype (64.2%), 32 had an NGS-based panel (47.8%), 10 had a region-specific MLPA® (MRC-Holland) (14.9%), 8 had a molecular karyotype (11.9%), 7 had *FMR1* gene testing (10.4%), and 4 had a region-specific FISH (6.0%).

Regarding clinical manifestations, most participants n = 35 (52.2%) presented with both MCA and NDD. A smaller proportion had IEI alone n = 3 (4.5%) or in combination with MCA n = 3 (4.5%) (Figure 1A). The number of affected systems ranged from 2 to 10, with a median of 6. Figure 1B shows the percentage of participants with each affected system. “Neurological” and “Head and Neck” were the most frequently affected systems, present in 54 cases (80.6%) of this second phase of the DECIPHERD project. Two mothers, one father, and one sibling presented at least one HPO term related to the patient’s phenotype and were therefore considered possibly affected.

**Figure 1.**
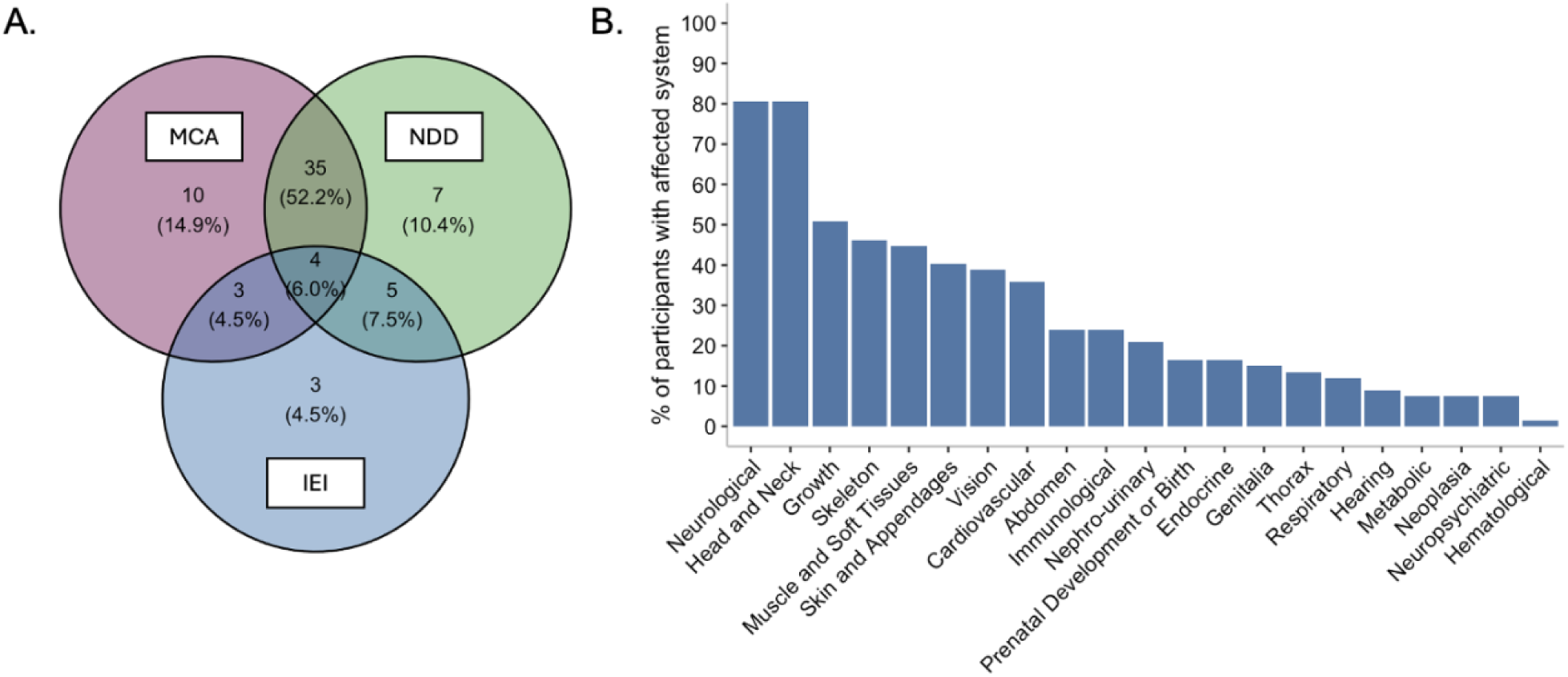
Inclusion Criteria and Clinical Manifestations. **A.** Number of participants meeting each inclusion criterion and their combinations. MCA: Multiple congenital anomalies. NDD: Neurodevelopmental disorders. IEI: Inborn errors of immunity (suspected). **B.** Percentage of participants with involvement of each system according to HPO’s hierarchy.

### Clinical variant interpretation results

In this second phase of the DECIPHERD project, we identified an informative result in 30 cases (44.8%). Of these, 23 cases were interpreted as categorical (34.3%) harboring a total of 31 variants, of which 19 (61.3%) had not been previously reported in ClinVar (26).

All cases had unique phenotypic and molecular findings. The clinical features and variants are described in Table 2 (Table 2 is presented at the end of the manuscript). In 17 of 23 of categorical cases (73.9%) we identified at least one P/LP variant in a gene associated with an autosomal dominant (AD) syndrome. Of this, 14 variants, corresponding to 12 cases (including two dual-diagnosis cases) were *de novo.* In four categorical cases (17.4%), we identified two P/LP variants in genes associated with autosomal recessive (AR) syndromes. Finally, in one case, we detected an LP variant associated with an X-linked recessive/dominant syndrome (4.3%), and in another, a dominant LP somatic variant in oral mucosa (4.3%). In one case, we found two additional VUS consistent with the patients’ phenotype, suggesting the possibility of more than one genetic diagnosis.

**Table 2:**
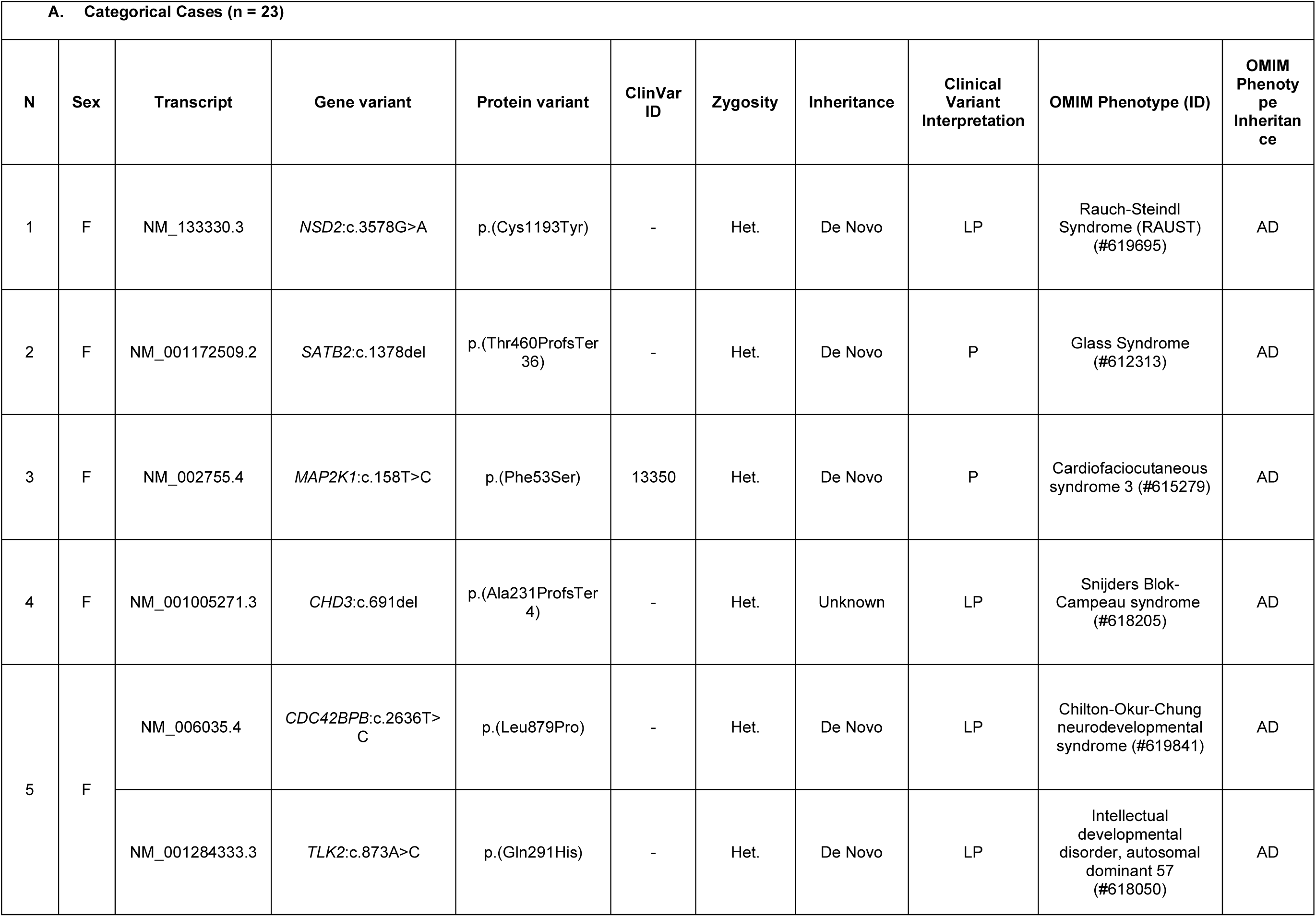
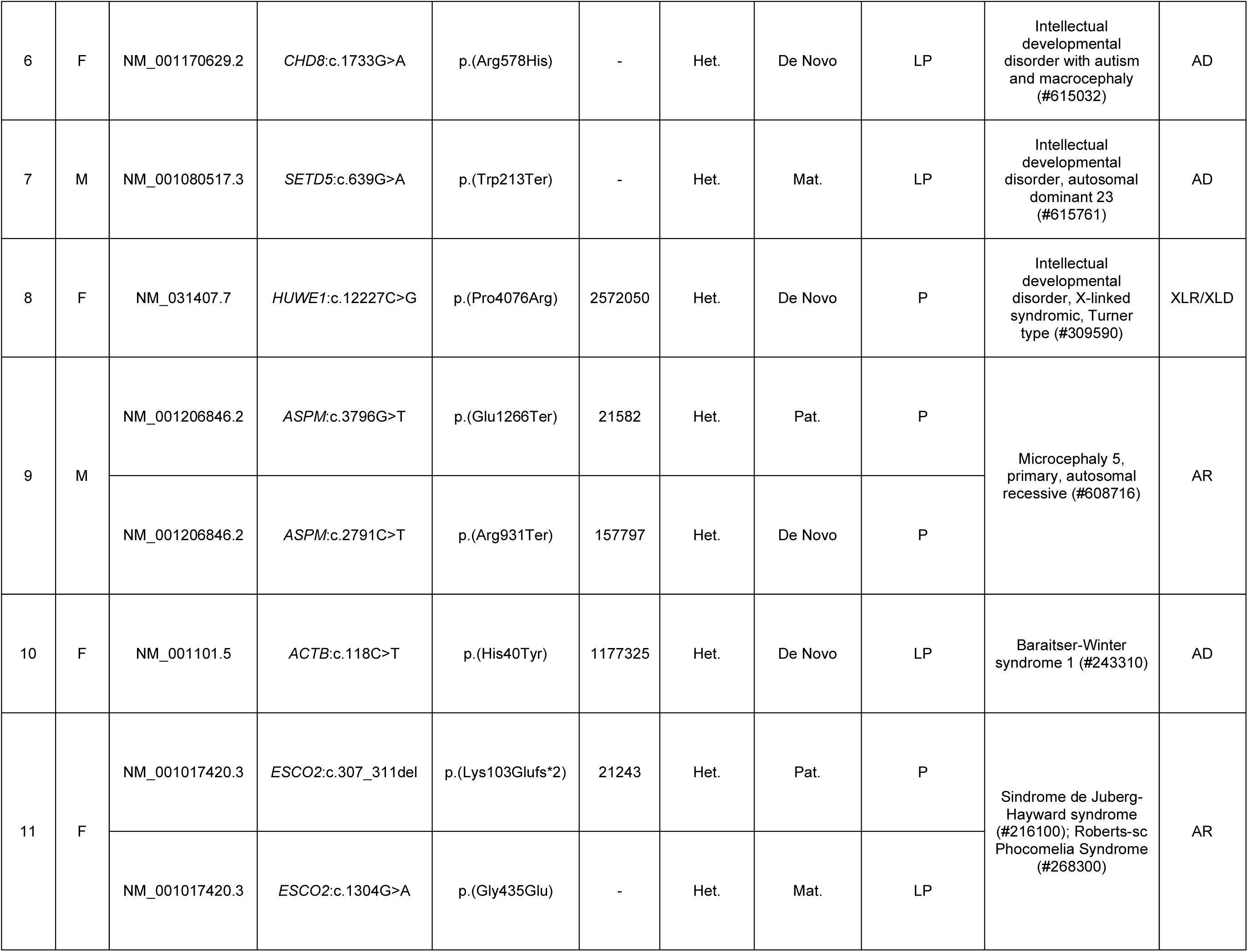
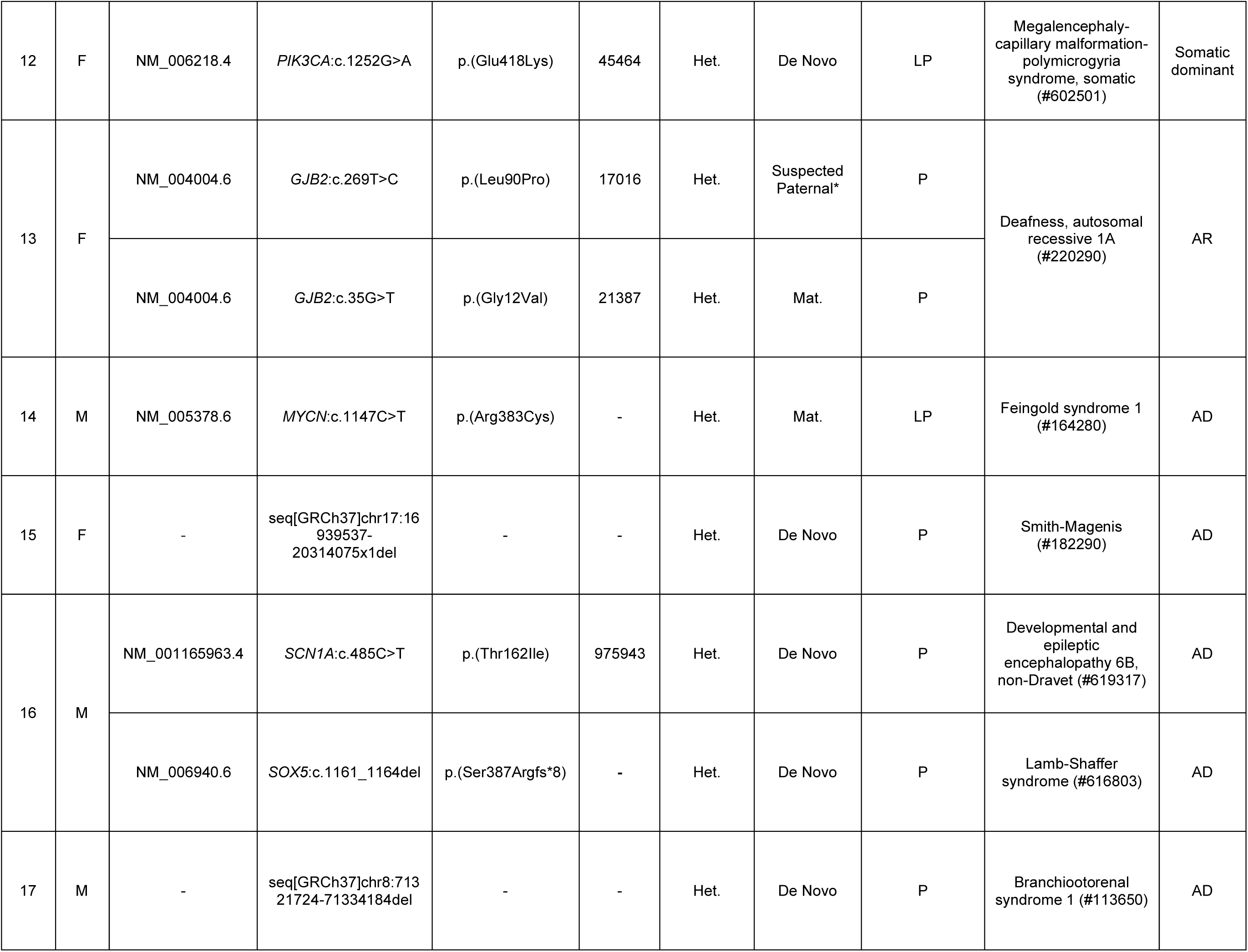
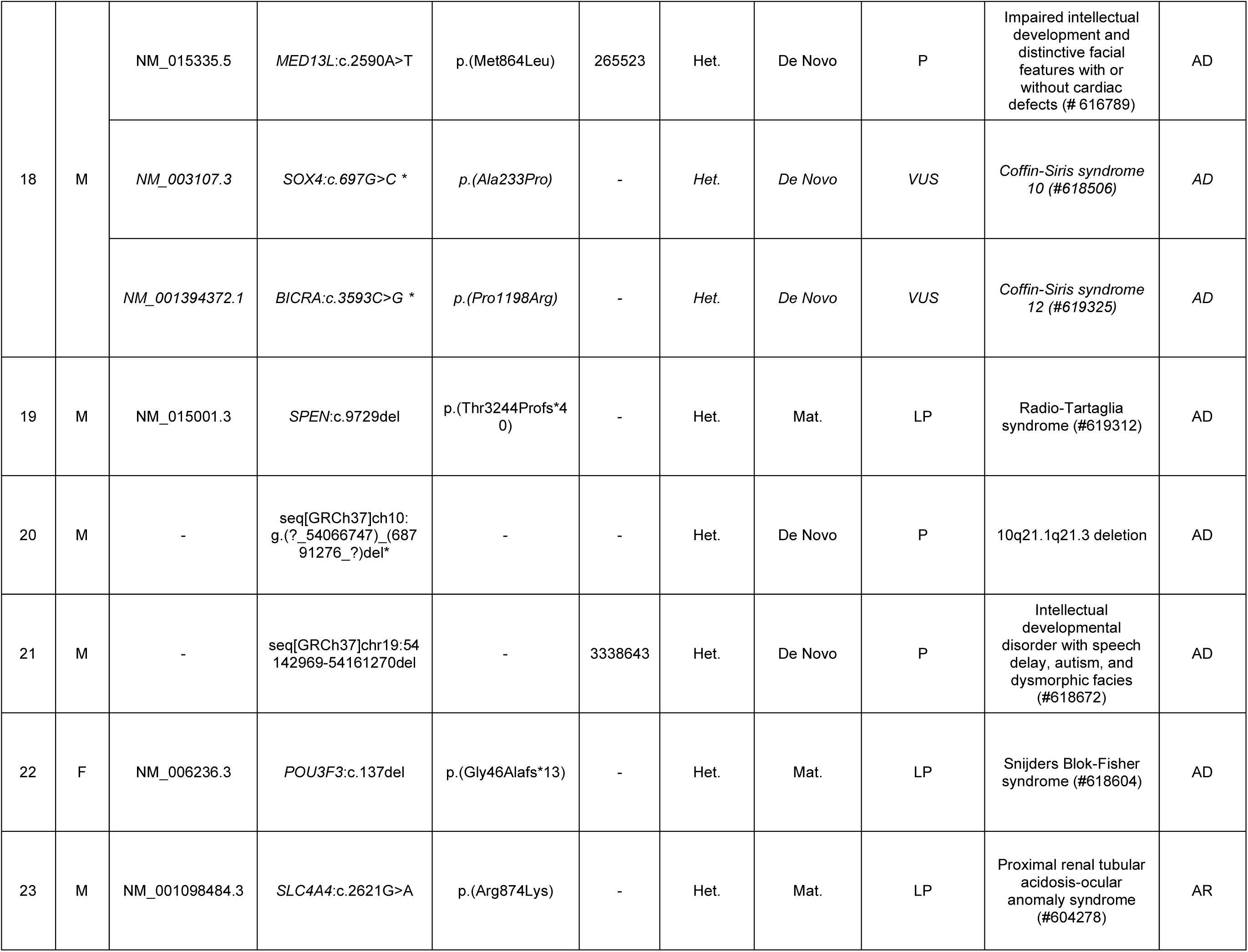
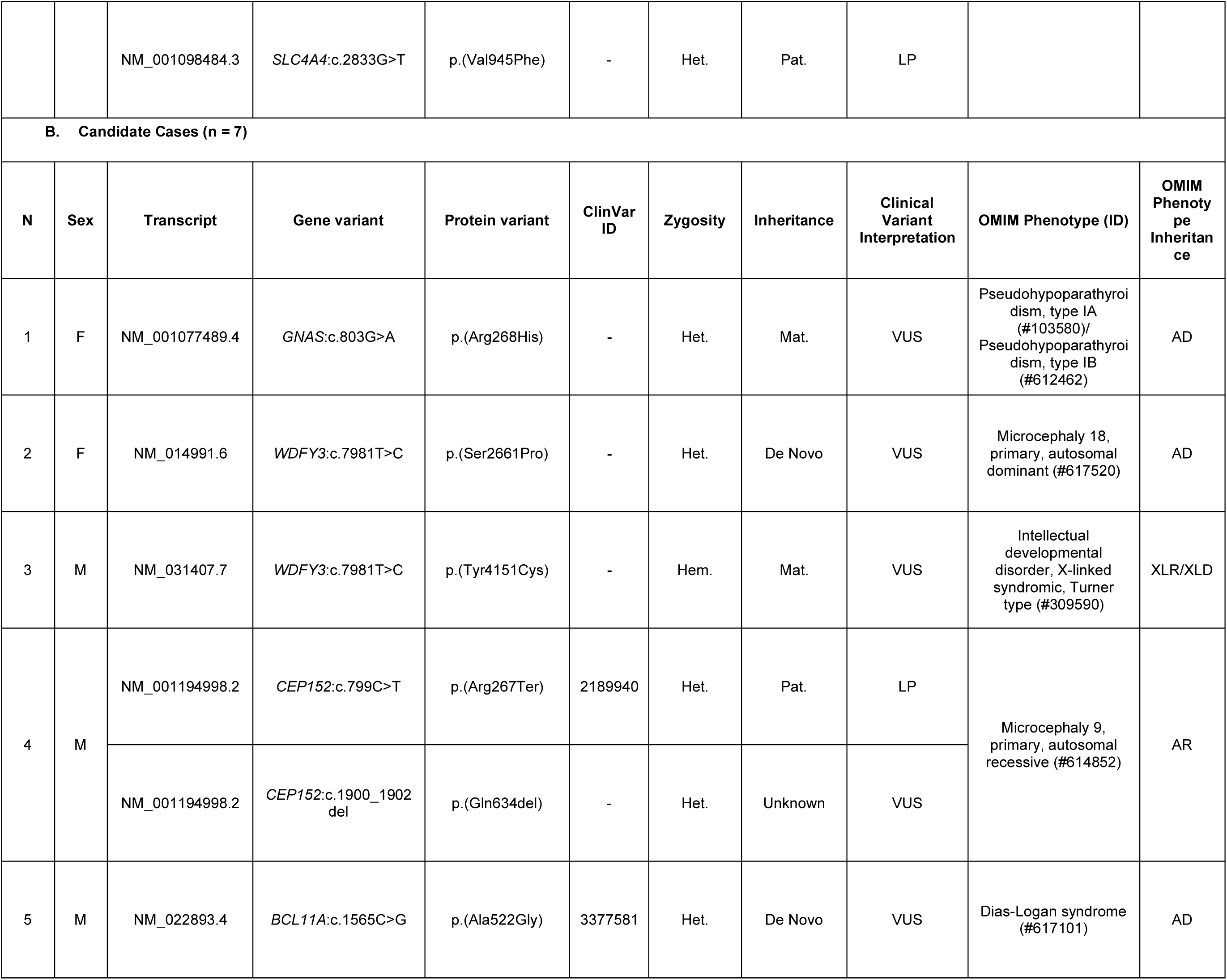
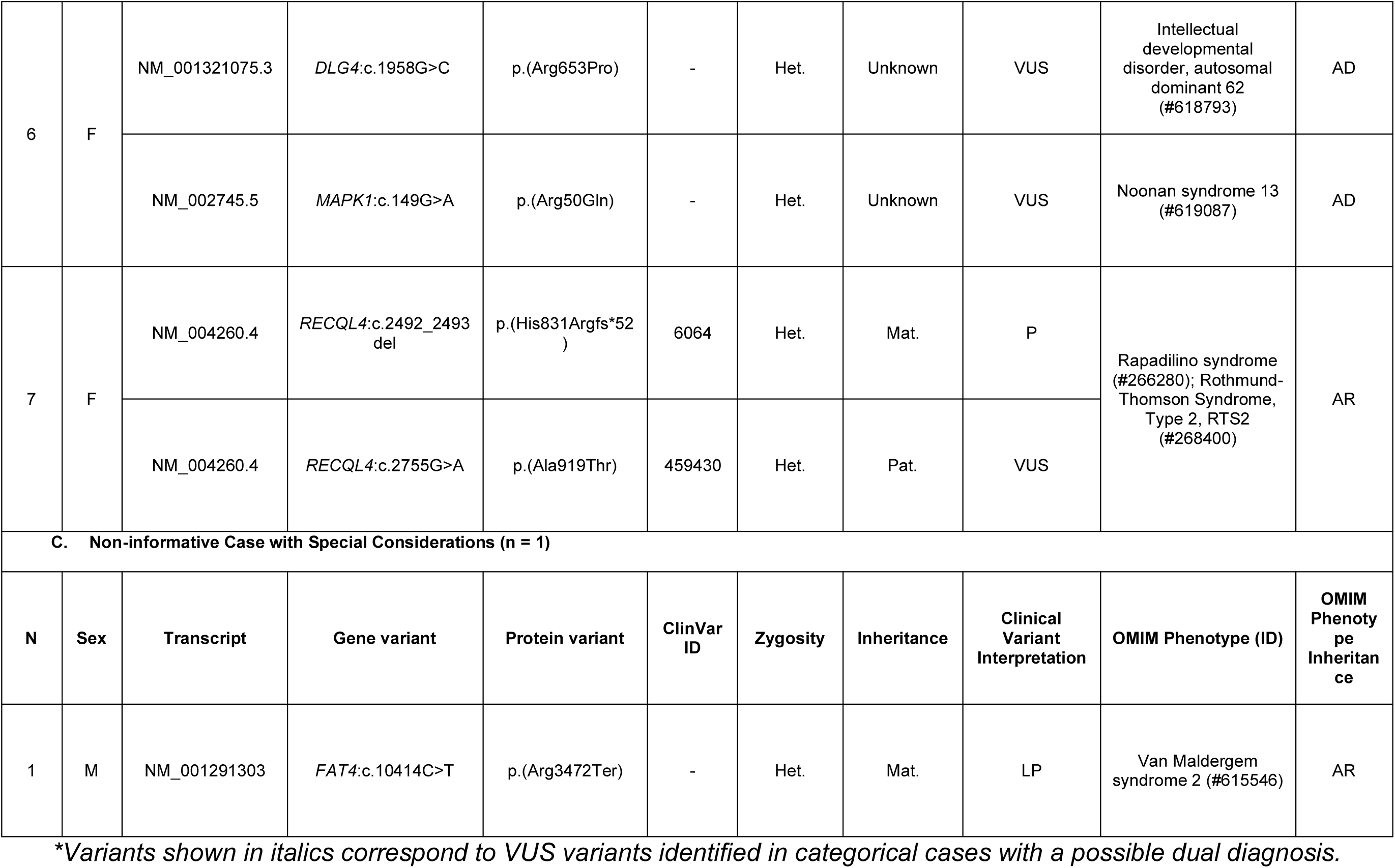
Cases with Identified Variants.

Regarding differences in classification, four cases initially categorized as candidate based on GeneSystem classification (June 2023 to January 2025) involving VUS in autosomal dominant (AD) syndromes, were subsequently reclassified as categorical cases, following the availability of new evidence through additional variant interpretation platforms (VarSome and Franklin) in October 2025. Similarly, one case, previously classified as a candidate case (VUS) was reclassified as non-informative to the variant being reclassified as likely benign. In the reclassification process, discrepant classifications between VarSome and Franklin were identified for five variants across 30 informative cases, affecting 16.6% of cases. The classification supported by the greater amount of evidence was retained.

Two of the 30 cases had a dual diagnosis. In the first case, a LP variant in the *CDC42BPB* gene, associated with AD Chilton-Okur-Chung neurodevelopmental syndrome (OMIM #619841), and a LP variant in the *TLK2* gene, associated with Intellectual developmental disorder, autosomal dominant 57 (OMIM #618050) were detected. Both variants were *de novo* and consistent with the patient’s phenotype, including intellectual disability and dysmorphic facial features. Although both syndromes have been reported to include ophthalmologic manifestations such as ptosis or strabismus, the more severe ophthalmological findings observed in this patient, specifically optic nerve aplasia, have not been previously described. The second dual diagnosis case had two pathogenic variants in the *SCN1A* and *SOX5* genes each, associated with Autosomal Dominant Developmental and epileptic encephalopathy 6B, non-Dravet (#619317) and Lamb-Shaffer syndrome (#616803) respectively, conditions that have overlapping neurodevelopmental features.

In addition, seven patients (10.4%) received a candidate result. Of these, four corresponded to cases with one or two VUS in genes associated with an AD syndrome consistent with the phenotype (two *de novo*, one maternally inherited and two of unknown parental origin in the same case). One case corresponded to a maternally inherited VUS in a gene associated with XL syndrome in a male participant. In two cases, we identified a LP or P variant and a VUS in genes associated with AR syndromes (Table 2).

No variants were identified in mitochondrial DNA in the four cases in whom a mitochondrial condition was suspected.

Among the non-informative cases, one presented potentially relevant findings that did not meet sufficient criteria to be considered informative, consisting of a single LP variant in the *FAT4* gene, associated with AR Van Maldergem syndrome (OMIM #615546), and concordant with patient’s phenotype.

### Factors associated with diagnostic yield

To assess factors associated with obtaining an informative result, we merged participants from the first (16) and second phases of the DECIPHERD project into a combined dataset (N = 167), given that they had similar ascertainment and analysis strategies and differed only in the sequencing and informatics platform used.

In terms of clinical presentation, we observed that the proportion of informative results was highest among participants presenting with both MCA and NDD (59%), whereas the lowest proportion was for patients with IEI alone (10%), as shown in Figure 2. Consistent with this distribution, bivariate regression analyses showed that the presence of MCA or NDD was associated with increased odds of obtaining informative results, with the stronger association observed when both conditions co-occurred (OR = 3.42, p-value = 0.0002). Additionally, presenting IEI, whether isolated or in combination with any of the other categories, was associated with lower odds of reaching a diagnosis (OR = 0.39, p-value = 0.015). The number of affected systems, sex, and age were not significant predictors of diagnostic outcome.

**Figure 2.**
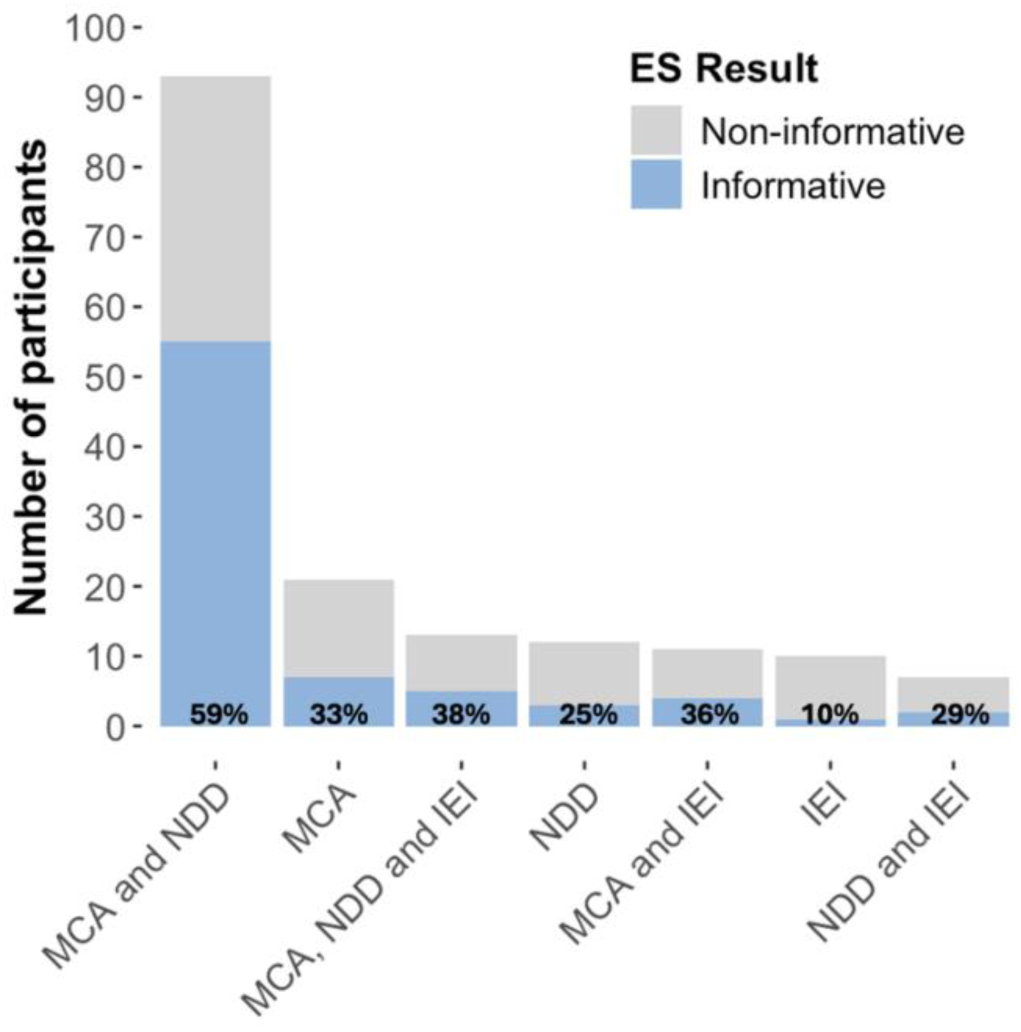
Diagnostic yield according to inclusion criteria. Bars represent the number of participants in each category. MCA: Multiple congenital anomalies. NDD: Neurodevelopmental disorder. IEI: Inborn errors of immunity.

Next, we evaluated whether prior genetic panel testing influenced the probability of receiving a diagnosis. In the combined dataset (first and second phase), 139 cases (83.2%) had undergone at least one previous non-informative genetic test, including 64 cases that had an NGS-based panel (38.3%). The results showed that having undergone a previous panel test was associated with a lower probability of reaching a diagnosis by ES (OR = 0.37, p-value = 0.003).

We also compared ES solo with duo and trio analysis, using ES solo as the reference category, and found no significant differences with the other two strategies (OR = 0.53, p-value = 0.378 and OR = 0.72, p-value = 0.543, respectively). Detailed results of the bivariate logistic regression analysis are provided in Table 3.

**Table 3:** Bivariate logistic regression analyses for factors associated with an informative diagnosis.

| Variable | Bivariate analysis |  |  |
| --- | --- | --- | --- |
|  | OR | [95%CI] | p |
| Sex (M) | 0.68 | 0.37 – 1.25 | 0.218 |
| Age | 1.00 | 0.97 – 1.04 | 0.911 |
| Presence of MCA (n = 138) * | 4.06 | 1.65 – 11.55 | **0.004** |
| Presence of NDD (n = 125) * | 2.70 | 1.30 – 5.95 | **0.010** |
| Presence of IEI (n = 41) * | 0.39 | 0.18 – 0.81 | **0.015** |
| Only NDD (n = 21) | 0.54 | 0.20 – 1.38 | 0.214 |
| Only MCA (n = 12) | 0.36 | 0.08 – 1.27 | 0.142 |
| Only IEI (n = 10) | 0.12 | 0.00 – 0.65 | **0.045** |
| NDD and MCA (n = 93) | 3.42 | 1.81 – 6.63 | **0.0002** |
| NDD, MCA and IEI (n = 13) | 0.71 | 0.21 – 2.23 | 0.566 |
| NDD and IEI (n = 11) | 0.65 | 0.16 – 2.24 | 0.505 |
| MCA and IEI (n = 7) | 0.45 | 0.06 – 2.17 | 0.353 |
| Affected Systems | 1.03 | 0.90 – 1.20 | 0.684 |
| Previous panel testing (n = 64) | 0.37 | 0.19 – 0.70 | **0.003** |
| ES duo (vs solo) | 0.53 | 0.11 – 2.11 | 0.378 |
| ES trio (vs solo) | 0.72 | 0.24 – 2.10 | 0.543 |

*\*With or without other criteria*

Regarding the platforms used, the proportion of informative cases was similar between the two versions of GeneSystems© platforms used in the second phase, GeneBytes® and GeneBytesX® (45.5% and 44.1%, respectively) with no significant differences (Chi-square, p-value = 1). Likewise, no significant differences were found when these platforms were compared with SOPHiA DDM software® (used in the first phase) (45.6%, Chi-square, p-value = 0.91).

Based on the analysis presented above, we subsequently developed a multivariate model including the variables that showed statistically significant associations, adjusted for sex and age, as shown in Figure 3. The presence of MCA or NDD was associated with higher odds of obtaining an informative diagnosis (MCA: OR = 2.97, p-value = 0.041; NDD: OR = 2.46, p-value = 0.036). In contrast, previous panel testing was associated with lower odds of an informative result (OR = 0.37, p-value = 0.005).

**Figure 3.**
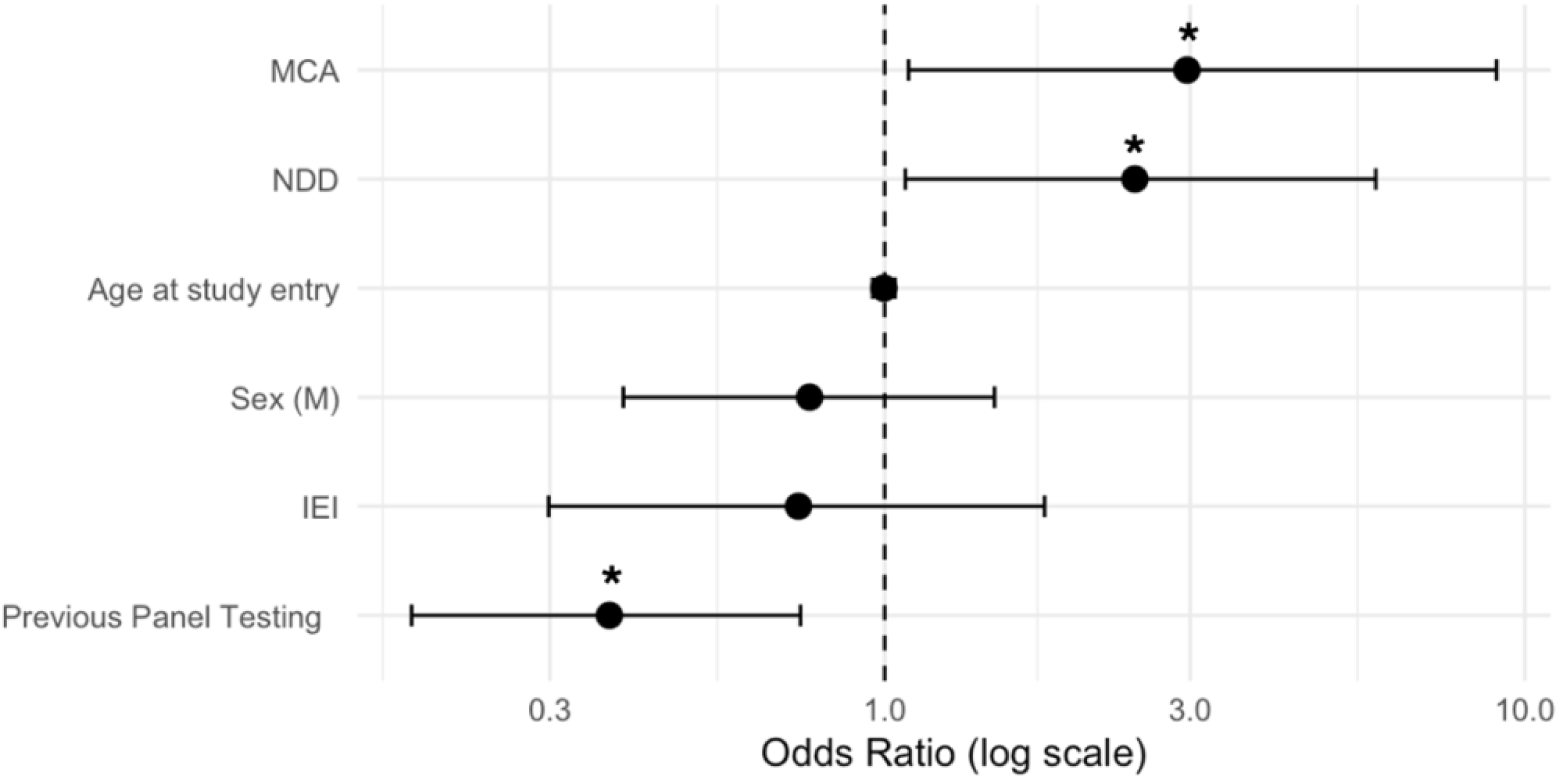
Results of the multivariate regression analysis. The figure shows the OR for each variable added to the regression model. An OR > 1 supports an increase in the chances of obtaining an informative result, whereas OR < 1 indicate the opposite. Asterisks denote statistical significance (p < 0.05). NDD: Neurodevelopmental disorder. MCA: Multiple congenital anomalies. IEI: Inborn errors of immunity.

Finally, we sought to evaluate a participant’s ancestry as a contributing factor to the diagnostic yield. We observed that the average proportion (± s.d.) of Native American ancestry in participants with an informative result was 0.54 ± 0.18, and with a non-informative result was 0.54 ± 0.12. We found no statistically significant differences between the groups, p-value = 0.974, Figure 4.

**Figure 4.**
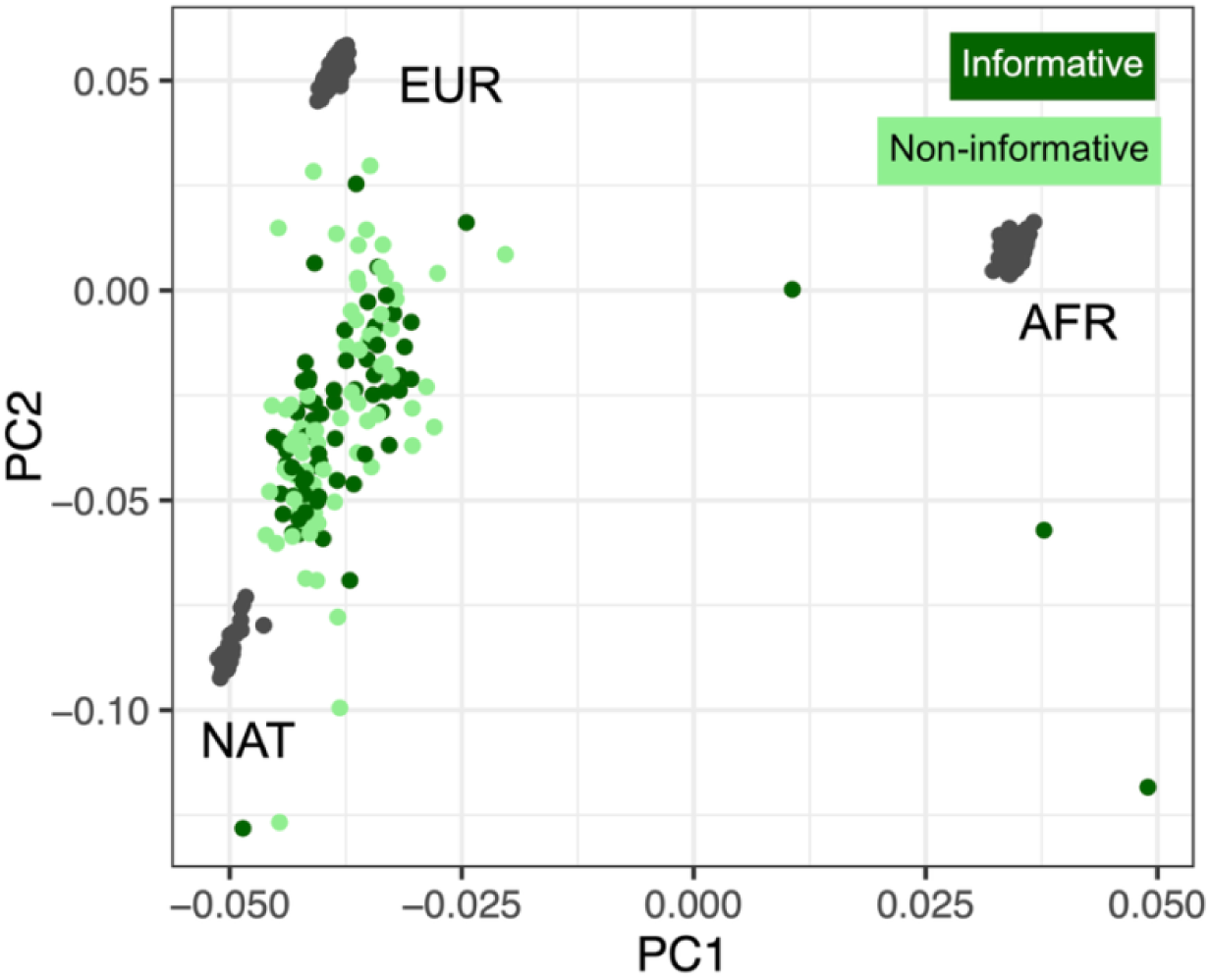
PCA projection of references and patients with and without informative findings. AFR: African, EUR: European, NAT: Native American.

## Discussion

Our study contributed to ending the diagnostic odyssey in a Chilean cohort of patients with RDs, confirming the utility of ES in different populations and heterogeneous clinical presentations. Similar to our initial study, the findings also contributed several previously unreported variants for known disorders, evidencing the value of including understudied populations in genomics research. Our results in this new phase of the study reached a diagnostic yield of 32.3%, comparable to that reported in the literature (1), including the first phase of the DECIPHERD project (30.1%) (16). This impacted a large number of patients who had already exhausted the genetic testing options available in Chile and had remained undiagnosed for a median time of almost a decade. Studies are underway to understand the personal and clinical implications of receiving a result.

Importantly, we characterized the demographic and clinical features associated with a higher probability of achieving a genetic diagnosis in this cohort. Considering the limited clinical genetic resources available in Chile, and that ES is not currently available or reimbursed in the health care system, our findings are key to guide clinical and public health decisions by prioritizing testing in individuals with MCA and NDD, that had the highest likelihood of receiving a diagnosis. The latter aligns with the clinical guidelines of the ACMG that recommend ES as a first- or second-tier test for individuals with congenital anomalies, developmental delays or intellectual disability (27). Patients presenting with IEI had a trend towards lower diagnostic yield, which could be due to other disease mechanisms that are not detectable with ES, such as mosaicism, environmental and multifactorial causes (28).

Most cases had undergone at least one previous genetic test, with a median of two and a range of two to four. This reflects the low availability of genetic tests in Chile compared with studies in other countries in which early use of ES or GS can replace numerous other tests (29). This situation may imply that cost-savings analysis may not be favorable towards ES in Chile, as seen in other countries such as Scotland (30)). Nevertheless, the potential added costs to the health system may bring other valuable benefits in terms of accelerating time to diagnosis, which in turn may reduce additional downstream testing and invasive procedures (31), enable earlier initiation of appropriate management and surveillance, inform reproductive planning and cascade testing (6,32), and deliver psychosocial benefits to patients and families by ending diagnostic uncertainty (33).

In our study, having prior negative panel test results decreased the likelihood of receiving an informative diagnosis by ES. This has been shown in other studies, in which the incremental diagnostic rate of ES or GS after a carefully selected phenotype-specific gene panel was low ((34). This finding could be due to selection biases towards more complex cases in our study, exhaustion of high yield genes, technical limitations of short read NGS used in both panels and exomes for the identification of complex structural variants or nucleotide repeat disorders, and lack of analysis of regulatory and other non-coding regions. Nevertheless, broader testing such as ES and GS have the added benefit of systematic reanalysis, shown to increase diagnosis by 10-15% after 1-2 years due to interval new discoveries, and thus produces durable genetic data that can be reinterpreted over time as medical knowledge advances (35,36).

Regarding the testing strategies, no differences were found between solo, duo and trio modalities. However, trio analysis facilitates the identification of *de novo* or recessive conditions, that accounted for most cases. Trio ES certainly increases sequencing costs, but decreases manual curation time, uncertain findings and need for segregation studies (Mordaunt et al., 2024; Tan et al., 2019). A detailed cost comparison study would provide key information for local implementation and coverage decisions. Of note, no difference was observed between the results with the different software used, reflecting the overall consistency of the platforms and our analysis strategy.

Ancestry analysis demonstrated a diagnostic yield in our admixed population that was comparable to that reported in the literature for individuals of predominantly European ancestry (4). However, unlike the mentioned study, Admixed American ancestry did not lower the diagnostic yield in our cohort. This is important because ancestry should not be considered a limitation to use ES.

We acknowledge limitations in our study, such as the relatively small sample size, but the results were consistent between both periods of the study, and with the published literature (1,2). In addition, since patients were presented from tertiary health care centers by clinicians aware of the study, the findings may not be generalizable to the broader undiagnosed patient population in the country.

In summary, this and our previous work show the feasibility and utility of ES for the diagnosis of URDs for patients in Chile. However, additional efforts are necessary to generate key evidence for local implementation and health system coverage, including cost-utility cost-benefit and other value-based analyses. The recent passing of the RD law in Chile (38) does not commit healthcare funding, but is a crucial step in the direction of the recognition of RDs and building a national plan.

Future work is also needed to identify a cause for non-informative cases. This will require data reanalysis and the use of additional strategies such as long read genome and transcriptome analysis. Similarly, advancing on solving the variants of unknown significance requires the improvement of analysis tools, data sharing representation of genome variation and accessible tools functional studies (39,40).

## Conclusion

In this study, we ended the diagnostic odyssey for a substantial number of patients with very heterogeneous clinical presentations and from an underrepresented population. As seen in the Chilean DECIPHERD cohort, most patients experienced prolonged diagnostic journeys and remained undiagnosed because ES and other tools are not routinely accessible and is preceded by multiple non-informative tests. Thus, while in genomic-inclusive health care, “undiagnosed” reflects the frontier of genomic discovery, in other settings, it largely reflects health-system inequities and delayed access to high-yield genomic technologies. Our work and others occurring in Latin America (Félix et al., 2022; Velasco et al., 2024) demonstrate the feasibility of incorporating these tools in the evaluation of patients. This is key to generating locally pertinent data that can be used to inform public policy decisions aimed at improving equitable access to technologies within health systems, that can shorten lengthy and inefficient diagnostic processes for patients living with URDs.

## Data Availability

The datasets used and/or analyses during the current study are available from the corresponding author on reasonable request.

## List of abbreviations

AFR: African
ES: Exome Sequencing
EUR: European
GS: Genome Sequencing
IEI: Inborn Errors of Immunity
NAT: Native American
NGS: Next Generation Sequencing
NND: Neurodevelopmental Disorders
MCA: Multiple Congenital Anomalies
RDs: Rare Diseases
UDRs: Undiagnosed Rare Diseases

## Declarations

### Ethics approval and consent to participate

The project was approved by the Research Ethics Committee at Facultad de Medicina, Clinica Alemana Universidad del Desarrollo (approval #2018-045) and all participants and/or legal guardians gave written informed consent.

### Consent for publication

Consent for publication was included in the informed consent form.

## Funding

ANID-Chile grants Fondecyt #1211411 (GMR), Fondecyt #11220642 and EQM150093 (BR-J), Fondecyt # 1221802 (CP), Fondecyt #11240332 (VF) and a donation from the Child Health Foundation, Birmingham, AL.

### Authors’ contributions

Conception and Design of the work: GMR, BR-J, CP

Data acquisition (Patient recruitment, clinical description) GMR, VF, LMM, CP, TH, FE, MJZ

Data analysis and interpretation. GMR, GM, BR-J, DB, GE LMM, CP, VF

Drafting and substantial revision of the manuscript GMR, GM, BR-J

All authors approved the submitted version

All authors agree to be both personally accountable for the author’s own contributions and to ensure that questions related to the accuracy or integrity of any part of the work, even ones in which the author was not personally involved, are appropriately investigated, resolved, and the resolution documented in the literature.

## Acknowledgements

To all participating patients and families

## REFERENCES

1. Chung CCY, Hue SPY, Ng NYT, Doong PHL, Chu ATW, Chung BHY. Meta-analysis of the diagnostic and clinical utility of exome and genome sequencing in pediatric and adult patients with rare diseases across diverse populations. Vol. 25, Genetics in Medicine. Elsevier B.V.; 2023.

2. Pandey R, Brennan NF, Trachana K, Katsandres S, Bodamer O, Belmont J, et al. A meta-analysis of diagnostic yield and clinical utility of genome and exome sequencing in pediatric rare and undiagnosed genetic diseases. Vol. 27, Genetics in Medicine. Elsevier B.V.; 2025.

3. Wright CF, Campbell P, Eberhardt RY, Aitken S, Perrett D, Brent S, et al. Genomic Diagnosis of Rare Pediatric Disease in the United Kingdom and Ireland. New England Journal of Medicine. 2023 Apr 27;388(17):1559–71.

4. Wojcik MH, Lemire G, Berger E, Zaki MS, Wissmann M, Win W, et al. Genome Sequencing for Diagnosing Rare Diseases. New England Journal of Medicine. 2024 Jun 6;390(21):1985–97.

5. Shan Z, Ding L, Zhu C, Sun R, Hong W. Medical care of rare and undiagnosed diseases: Prospects and challenges. Vol. 2, Fundamental Research. KeAi Communications Co.; 2022. p. 851–8.

6. Malinowski J, Miller DT, Demmer L, Gannon J, Maria Pereira E, Schroeder MC, et al. Systematic evidence-based review: outcomes from exome and genome sequencing for pediatric patients with congenital anomalies or intellectual disability. 2020; Available from: 10.1038/s41436-

7. Boycott KM, Rath A, Chong JX, Hartley T, Alkuraya FS, Baynam G, et al. International Cooperation to Enable the Diagnosis of All Rare Genetic Diseases. Vol. 100, American Journal of Human Genetics. Cell Press; 2017. p. 695–705.

8. Boycott KM, Azzariti DR, Hamosh A, Rehm HL. Seven years since the launch of the Matchmaker Exchange: The evolution of genomic matchmaking. Hum Mutat. 2022 Jun 1;43(6):659–67.

9. Velasco HM, Bertoli-Avella A, Jaramillo CJ, Cardona DS, González LA, Vanegas MN, et al. Facing the challenges to shorten the diagnostic odyssey: first Whole Genome Sequencing experience of a Colombian cohort with suspected rare diseases. European Journal of Human Genetics. 2024 Oct 1;32(10):1327–37.

10. Encina G, Castillo-Laborde C, Lecaros JA, Dubois-Camacho K, Calderón JF, Aguilera X, et al. Rare diseases in Chile: Challenges and recommendations in universal health coverage context. Orphanet J Rare Dis. 2019 Dec 11;14(1).

11. Lawrence MG, Rider NL, Cunningham-Rundles C, Poli MC. Disparities in Diagnosis, Access to Specialist Care, and Treatment for Inborn Errors of Immunity. Journal of Allergy and Clinical Immunology: In Practice. 2024 Feb 1;12(2):282–7.

12. Wainstock D, Katz A. Advancing rare disease policy in Latin America: a call to action. Vol. 18, The Lancet Regional Health - Americas. Elsevier Ltd; 2023.

13. Department of Health D and A. https://www.msac.gov.au/applications/1476#application-details. 2026. Application 1476 – Genetic testing for childhood syndromes.

14. El Chehadeh S, Heide S, Quélin C, Rio M, Margot H, Geneviève D, et al. Genome sequencing for the diagnosis of intellectual disability as a paradigm for rare diseases in the French healthcare setting: the prospective DEFIDIAG study. Genome Med. 2025 Dec 1;17(1).

15. The 100000 Genomes Project Pilot Investigators. 100,000 Genomes Pilot on Rare Disease Diagnosis in Health Care — Preliminary Report. 2021;

16. Poli MC, Rebolledo-Jaramillo B, Lagos C, Orellana J, Moreno G, Martín LM, et al. Decoding complex inherited phenotypes in rare disorders: the DECIPHERD initiative for rare undiagnosed diseases in Chile. European Journal of Human Genetics. 2024 Oct 1;32(10):1227–37.

17. Gargano MA, Matentzoglu N, Coleman B, Addo-Lartey EB, Anagnostopoulos A V., Anderton J, et al. The Human Phenotype Ontology in 2024: phenotypes around the world. Nucleic Acids Res. 2024 Jan 5;52(D1):D1333–46.

18. Harris PA, Taylor R, Minor BL, Elliott V, Fernandez M, O’Neal L, et al. The REDCap consortium: Building an international community of software platform partners. Vol. 95, Journal of Biomedical Informatics. Academic Press Inc.; 2019.

19. Rehm HL, Berg JS, Brooks LD, Bustamante CD, Evans JP, Landrum MJ, et al. ClinGen-The Clinical Genome Resource. Vol. 23, n engl j med. 2015.

20. Foreman J, Perrett D, Mazaika E, Hunt SE, Ware JS, Firth HV. DECIPHER: Improving Genetic Diagnosis Through Dynamic Integration of Genomic and Clinical Data. 2025;27:8. Available from: 10.1146/annurev-genom-102822-

21. Genoox. Franklin by Genoox. 2025.

22. Kopanos C, Tsiolkas V, Kouris A, Chapple CE, Albarca Aguilera M, Meyer R, et al. VarSome: the human genomic variant search engine. Bioinformatics. 2019 Jun 1;35(11):1978–80.

23. Tavtigian S V., Harrison SM, Boucher KM, Biesecker LG. Fitting a naturally scaled point system to the ACMG/AMP variant classification guidelines. Hum Mutat. 2020 Oct 1;41(10):1734–7.

24. Alexander DH, Novembre J, Lange K. Fast model-based estimation of ancestry in unrelated individuals. Genome Res. 2009;19(9).

25. R Core Team. R: A language and environment for statistical computing. Vienna, Austria; 2021.

26. National Center for Biotechnology Information. https://www.ncbi.nlm.nih.gov/clinvar/. 2026. ClinVar.

27. Manickam K, McClain MR, Demmer LA, Biswas S, Kearney HM, Malinowski J, et al. Exome and genome sequencing for pediatric patients with congenital anomalies or intellectual disability: an evidence-based clinical guideline of the American College of Medical Genetics and Genomics (ACMG). Genetics in Medicine. 2021;23(11).

28. Poli MC, Aksentijevich I, Bousfiha AA, Cunningham-Rundles C, Hambleton S, Klein C, et al. Human inborn errors of immunity: 2024 update on the classification from the International Union of Immunological Societies Expert Committee. Journal of Human Immunity. 2025 May 5;1(1).

29. Stark Z, Schofield D, Alam K, Wilson W, Mupfeki N, Macciocca I, et al. Prospective comparison of the cost-effectiveness of clinical whole-exome sequencing with that of usual care overwhelmingly supports early use and reimbursement. Genetics in Medicine. 2017;19(8).

30. Abbott M, Ryan M, Hernández R, McKenzie L, Heidenreich S, Hocking L, et al. Should Scotland provide genome-wide sequencing for the diagnosis of rare developmental disorders? A cost-effectiveness analysis. European Journal of Health Economics. 2025 Apr 1;26(3):503–12.

31. Yeung A, Tan NB, Tan TY, Stark Z, Brown N, Hunter MF, et al. A cost-effectiveness analysis of genomic sequencing in a prospective versus historical cohort of complex pediatric patients. Genetics in Medicine. 2020;22(12).

32. Shickh S, Mighton C, Uleryk E, Pechlivanoglou P, Bombard Y. The clinical utility of exome and genome sequencing across clinical indications: a systematic review. Vol. 140, Human Genetics. Springer Science and Business Media Deutschland GmbH; 2021. p. 1403–16.

33. Cabieses B, Obach A, Roberts A, Repetto G. Therapeutic trajectories of families with rare diseases in Chile from the perspectives of patients, carers, and healthcare workers: a qualitative study. Orphanet Journal of Rare Diseases. 2025 Dec 1;20(1).

34. Wilke MVMB, Klee EW, Dhamija R, Fervenza FC, Thomas B, Leung N, et al. Diagnostic yield of exome and genome sequencing after non-diagnostic multi-gene panels in patients with single-system diseases. Orphanet Journal of Rare Diseases. 2024 Dec 1;19(1).

35. Dai P, Honda A, Ewans L, McGaughran J, Burnett L, Law M, et al. Recommendations for next generation sequencing data reanalysis of unsolved cases with suspected Mendelian disorders: A systematic review and meta-analysis. Vol. 24, Genetics in Medicine. Elsevier B.V.; 2022. p. 1618–29.

36. Best S, Fehlberg Z, Richards C, Quinn MCJ, Lunke S, Spurdle AB, et al. Reanalysis of genomic data in rare disease: current practice and attitudes among Australian clinical and laboratory genetics services. European Journal of Human Genetics. 2024 Nov 1;32(11):1428–35.

37. Mordaunt DA, Gonzalez FS, Lunke S, Eggers S, Sadedin S, Chong B, et al. The cost of proband and trio exome and genome analysis in rare disease: A micro-costing study. Genetics in Medicine. 2024 Apr 1;26(4).

38. MINISTERIO DE SALUD. Ley de Enfermedades Poco Frecuentes, Raras o Huérfanas. 21.743 Chile: https://www.bcn.cl/leychile/navegar?idNorma=1213122; 2025.

39. Wojcik MH, Reuter CM, Marwaha S, Mahmoud M, Duyzend MH, Barseghyan H, et al. Beyond the exome: What’s next in diagnostic testing for Mendelian conditions. Vol. 110, American Journal of Human Genetics. 2023.

40. Fowler DM, Rehm HL. Will variants of uncertain significance still exist in 2030? Vol. 111, American Journal of Human Genetics. Cell Press; 2024. p. 5–10.

41. Félix TM, de Oliveira BM, Artifon M, Carvalho I, Bernardi FA, Schwartz I V.D., et al. Epidemiology of rare diseases in Brazil: protocol of the Brazilian Rare Diseases Network (RARAS-BRDN). Orphanet J Rare Dis. 2022 Dec 1;17(1).

